# A Changing Landscape of Carbapenem-Resistant *Escherichia coli* in Hong Kong: Emergence of *bla*_NDM-5_-Carrying ST69 across Clinical and Food Sources

**DOI:** 10.64898/2026.08.14.26360231

**Authors:** Iain Chi-Fung Ng, Ivan Tak-Fai Wong, Jake Siu-Lun Leung, Lam-Kwong Lee, Alison Yee-Ting Lam, Ho-Ching Tong, Shue-Kei Chan, Choi-Ying Wong, Annie Wing-Tung Lee, Wing-Yin Tam, Jia-Ying Zhang, Edward M Hill, Mei-Fan Hung, Miranda Chong-Yee Yau, River Chun-Wai Wong, Jason Chi-Ka Cheng, Cindy Wing-Sze Tse, Jimmy Yiu-Wing Lam, Viola Chi Ying Chow, Sandy Ka-Yee Chau, Franklin Wang-Ngai Chow, Polly Hang Mei Leung, Gilman Kit-Hang Siu

**Author notes:** These authors contributed equally to this work (Iain Chi-Fung Ng, Ivan Tak-Fai Wong). These authors are co-corresponding authors (Franklin Wang-Ngai Chow, Polly Hang Mei Leung, Gilman Kit-Hang Siu). Correspondence to: Franklin Wang-Ngai CHOW, Polly Hang Mei LEUNG, Gilman Kit-Hang SIU, Department of Health Technology and Informatics, Hong Kong Polytechnic University, Hong Kong.

## Abstract

Carbapenem-resistant *Escherichia coli* (CR-*E. coli*) is an emerging One Health threat, but recent shifts in predominant lineages and genomic links between clinical and food reservoirs in Hong Kong remain poorly defined. We analyzed 271 CR-*E. coli* isolates from four hospitals (2022-2026) and 585 isolates recovered from 4,917 retail food samples (2022-2025). Isolates underwent antimicrobial susceptibility testing, whole-genome sequencing, multilocus sequence typing, resistance-gene and plasmid profiling, core-genome SNP phylogenetics, and comparative genomics. Food isolates were mainly from raw pork (268/585, 45.8%) and raw chicken (231/585, 39.5%). *bla*_NDM-5_ was detected in 527/585 (90.1%) food and 241/271 (88.9%) clinical isolates. ST69 was the most frequent defined sequence type in both collections, representing 44/585 (7.5%) food and 36/271 (13.3%) clinical isolates, in contrast to the heterogeneous lineages and carbapenemases previously reported in Hong Kong. Applying a predefined ≤50-pairwise-SNP threshold for close genomic relatedness, core-genome phylogeny of 80 ST69 isolates identified two major mixed-source clusters collectively comprising 28 clinical and 27 food isolates. Clustered isolates showed similar antimicrobial resistance profiles, carried *bla*_NDM-5_ and *bla*_TEM-1_, and were associated with IncI1 MLST | ST136 plasmids. Comparative analyses showed >99.85% average nucleotide identity and broad conservation of the *bla*_NDM-5_-associated plasmid backbone across sources. These findings indicate the emergence of *bla*_NDM-5_-carrying ST69 as a prominent CR-*E. coli* lineage in Hong Kong and demonstrate close genomic relatedness between selected clinical and retail food isolates. Although transmission direction have not been inferred yet, the findings support integrated One Health surveillance and source-tracing across clinical, food, animal, and environmental sectors.

## 1. Introduction

Carbapenem-resistant *Enterobacterales* (CRE) constitute a major threat to clinical care and public health. Carbapenems are important last-line agents for treating severe infections caused by multidrug-resistant Gram-negative bacteria. The emergence of CRE substantially limits therapeutic options. Invasive CRE infections are associated with prolonged hospitalization, increased healthcare costs, and mortality rates exceeding 30–40% in some patient populations [1–4]. The spread of carbapenem resistance is facilitated by mobile genetic elements, including conjugative plasmids, transposons, and integrons, which enable carbapenemase genes to move within and between bacterial species, driving widespread dissemination [3–6]. Reflecting the urgency of this threat, the World Health Organization has classified carbapenem-resistant *Enterobacterales* as bacterial pathogens of critical public health priority [1].

Within this group, carbapenem-resistant *Escherichia coli* (CR-*E. coli*) warrants particular attention because *E. coli* is a leading cause of urinary tract, bloodstream, intra-abdominal, and other community- and healthcare-associated infections [7–9]. Evidence from China and the surrounding region indicates that CR-*E. coli* is of increasing clinical importance because of its sustained circulation, considerable clonal diversity, and changing resistance mechanisms over time [10–12]. In Hong Kong, surveillance by the Centre for Health Protection similarly showed an increase in *E. coli* isolates with reduced susceptibility to carbapenems in Hospital Authority general acute hospitals, rising from 209 of 61,799 isolates (0.3%) in 2021 to 744 of 65,729 isolates (1.1%) in 2024 [13]. These observations highlight the growing clinical relevance of carbapenem resistance in *E. coli* and the need for contemporary genomic surveillance.

Beyond its role as a human pathogen, *E. coli* functions as a ubiquitous intestinal commensal in humans and animals. Its ability to persist across diverse ecological niches allows it to act as a reservoir and vehicle for antimicrobial-resistance determinants at the human–animal–food– environment interface [8,14,15].The increasing detection of carbapenemase-producing *Enterobacterales* (CPE) in the food sector represents a parallel concern. Antimicrobial-resistance surveillance of raw meat in Hong Kong showed that the proportion of samples positive for CPE increased from 81/851 (9.5%) between 2019 and 2020 to 123/584 (21.1%) in 2022, 75/316 (23.7%) in 2023, and 185/677 (27.3%) in 2024 [16]. Studies from mainland China have likewise identified carbapenem-resistant *E. coli* and other carbapenem-resistant *Enterobacterales* in food-producing animals, porcine fecal samples, farm-associated environments, and retail meat. Genomic investigations have further demonstrated the dissemination of *bla*_NDM-5_-bearing IncX3 plasmids among isolates from food animals and retail meat [15,17,18]. The increasing occurrence of clinically relevant resistance determinants in animal-derived foods suggests that the food chain may serve as an important exposure interface and contribute to their wider ecological circulation. These observations provide a strong rationale for investigating CR-*E. coli* within a One Health framework, while recognizing that genomic similarity may reflect dissemination or acquisition from a common upstream source.

Despite the growing public health importance of CR-*E. coli* in Hong Kong, contemporary information on its genomic epidemiology remains limited. A previous longitudinal study of clinical isolates from 2011 to 2017 revealed a heterogeneous population structure, with ST405, ST131, and ST48 (each 6/78, 7.7%), alongside ST542 (5/78, 6.4%), as the predominant sequence types [19]. Across the entire clinical cohort, *bla*_NDM-5_ (39/78, 50.0%) represented one component of a diverse carbapenemase profile that co-circulated alongside *bla*_IMP-4_ (17/78, 21.8%), *bla*_NDM-1_ (10/78, 12.8%), and *bla*_OXA-48-like_ (9/78, 11.5%) [19]. Consequently, whether local clinical lineages and resistance determinants have shifted over time remains uncharacterized. Furthermore, the high-resolution genetic relatedness and plasmid exchange between contemporary clinical and food-derived CR-*E. coli* populations remain understudied.

To address these knowledge gaps, we investigated CR-*E. coli* isolates recovered from clinical specimens collected at four Hong Kong hospitals between January 2022 and March 2026 and from retail food samples collected throughout Hong Kong between January 2022 and December 2025. Our objectives were to define the contemporary molecular epidemiology of CR-*E. coli* in Hong Kong, assess changes relative to previously reported local patterns, and determine the degree of genomic relatedness between clinical and food-derived isolates. Through this One Health approach, we sought to evaluate cross-sectoral co-circulation and shared genomic features across clinical and food sectors.

## 2. Materials and Methods

### 2.1. Clinical isolate collection

A total of 271 clinical isolates of CR-*E. coli* were collected from four hospitals in Hong Kong between January 2022 and March 2026, namely Kwong Wah Hospital, Pamela Youde Nethersole Eastern Hospital, Prince of Wales Hospital, and United Christian Hospital. The isolates were recovered from a range of clinical specimens, including rectal swabs (n=212), urine (n=29), stool (n=17), blood (n=4), wound swabs (n=4), and other specimen types (n=5) (Figure 1).

**Figure 1:**
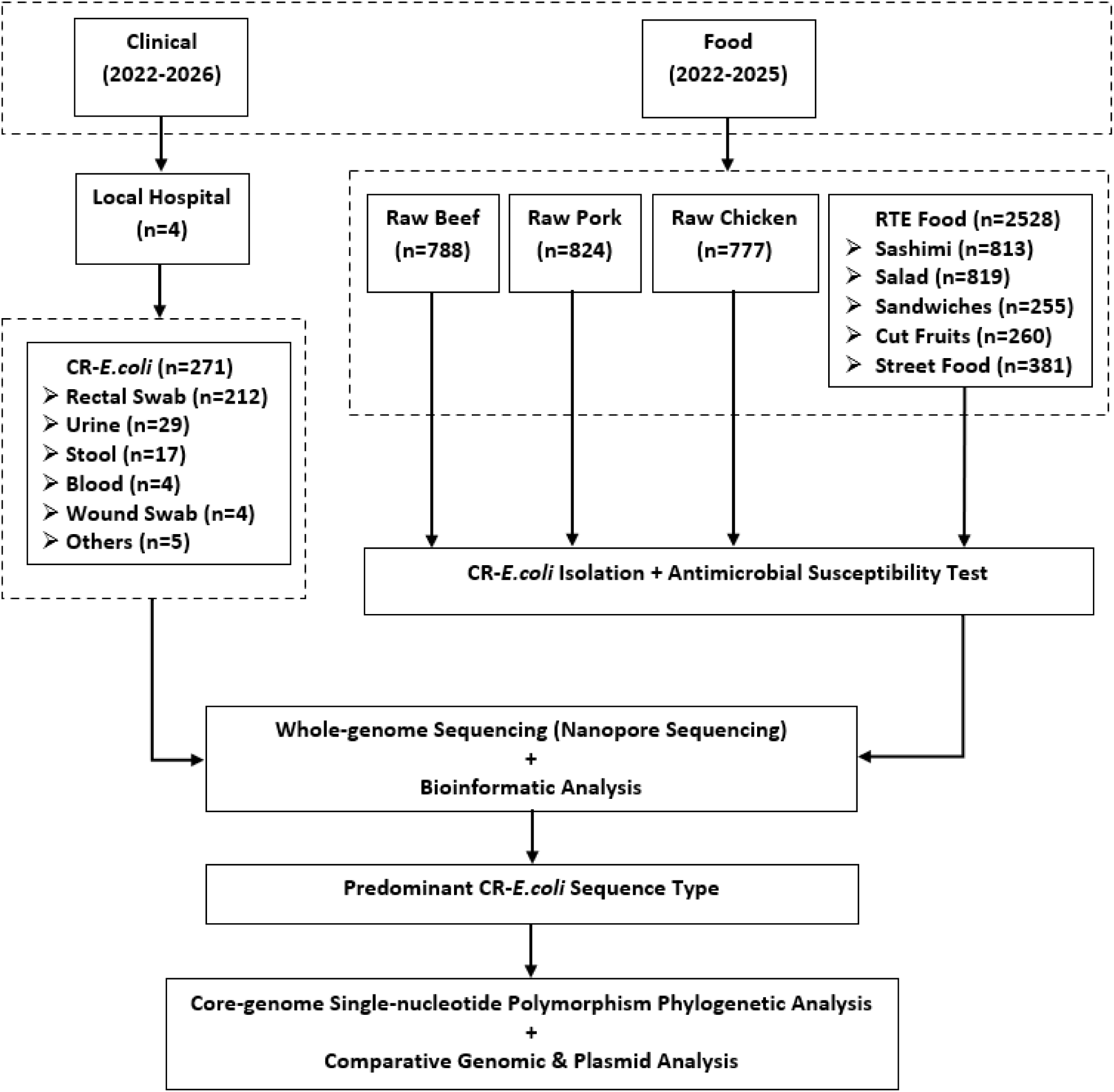
Overview of the study design and analytical workflow. A total of 585 carbapenem-resistant *Escherichia coli* isolates were recovered from 4,917 retail food samples collected across Hong Kong during 2022–2025, while 271 clinical isolates were collected from four hospitals during 2022–2026. All isolates underwent species identification, antimicrobial susceptibility testing, whole-genome sequencing, resistance-gene detection, and plasmid typing. Isolates belonging to the predominant shared sequence type were selected for core-genome SNP phylogenetic, comparative genomic, and *bla*_NDM-5_-associated plasmid analyses.

### 2.2. Food sample collection and processing

Between January 2022 and December 2025, a total of 4,917 food samples were systematically collected on a weekly basis from supermarkets, retail shops, and wet markets across all districts of Hong Kong. The collected samples comprised raw meat (n=2,389) and ready-to-eat (RTE) foods (n=2,528). Specifically, the raw meat category consisted of raw pork (n=824), raw beef (n=788), and raw chicken (n=777). The RTE food category comprised sashimi (n=813), salad (n=819), cut fruit (n=260), sandwiches (n=255), and street food (n=381) (Figure 1).

Sample processing and pre-enrichment were conducted in accordance with the European Union Reference Laboratory for Antimicrobial Resistance protocol for isolating ESBL-, AmpC-, and carbapenemase-producing *Escherichia coli* from fresh meat [20]. For each sample, 25 g of food was homogenized in 225 mL of sterile buffered peptone water (BPW; Oxoid CM1049, UK) in a stomacher bag using a stomacher blender (BagMixer 400P, Interscience, France) at 230 rpm for 90 s. The homogenates were then incubated at 35–37 °C for 24 h.

Following pre-enrichment, 0.5 mL of each homogenate was transferred into 4.5 mL of brain heart infusion (BHI) broth (Oxoid, UK) supplemented with vancomycin (10 μg/mL; Macklin, China) and meropenem (0.5 μg/mL; Macklin, China), followed by incubation at 35–37°C for 24 h. Subsequently, 10 μL of the enriched broth was streaked onto CHROMagar mSuperCARBA agar (CHROMagar, France) and incubated at 35–37°C for 24 h. Presumptive CR-*E. coli* isolates were screened and selected based on the manufacturer’s instructions, specifically identifying colonies that displayed a dark pink to reddish color.

### 2.3. Isolate identification and antimicrobial susceptibility testing

To confirm the species and the carbapenem-resistant phenotype, all presumptive CR-*E. coli* isolates were first subcultured to obtain pure colonies. Species identification was performed by matrix-assisted laser desorption/ionization time-of-flight mass spectrometry (MALDI-TOF MS) using a Microflex LT instrument (Bruker Daltonics, Germany). Spectral profiles were analysed using the MALDI Biotyper system (MALDI Biotyper Sirius one, Bruker Daltonics; MBT Compass Library-Version 12.0.0.0-[REF:1829023]) according to the manufacturer’s instructions.

Following species confirmation, antimicrobial susceptibility testing (AST) was conducted on the pure *E. coli* isolates to confirm carbapenem resistance and determine their broader susceptibility profiles. AST was performed using the Kirby–Bauer disk diffusion method on Mueller–Hinton agar in accordance with the Clinical and Laboratory Standards Institute (CLSI) Performance Standards for Antimicrobial Susceptibility Testing (M100) [21]. Bacterial suspensions were prepared directly from the pure colonies and adjusted to a 0.5 McFarland standard. Inoculated plates were incubated at 35 ± 2 °C for 16–18 h in ambient air, and inhibition zone diameters were measured and interpreted according to CLSI M100 breakpoints [21].

The antimicrobial agents tested by disk diffusion included carbapenems (imipenem 10 μg, meropenem 10 μg, doripenem 10 μg, ertapenem 10 μg), cephalosporins (cefepime 30 μg, cefotaxime 30 μg, cefoxitin 30 μg, ceftazidime 30 μg, ceftriaxone 30 μg), penicillins (ampicillin 10 μg), fluoroquinolones (ciprofloxacin 5 μg, nalidixic acid 30 μg), aminoglycosides (gentamicin 10 μg), tetracyclines (tetracycline 30 μg), phenicols (chloramphenicol 30 μg), and folate pathway antagonists (trimethoprim/sulfamethoxazole 25 μg).

### 2.4. Whole-genome sequencing and bioinformatic analysis

Genomic DNA was extracted from cultured CR-*E. coli* isolates using the QIAamp BiOstic Bacteremia DNA Kit (Qiagen, Germany) according to the manufacturer’s instructions. Whole-genome sequencing (WGS) was performed on a GridION Mk1 platform (Oxford Nanopore Technologies, UK). Sequencing libraries were prepared using the Rapid Barcoding Kit (SQK-RBK114.96; Oxford Nanopore Technologies, UK) following the manufacturer’s protocol. A normalized library input of 800 ng was loaded onto an R10.4.1 flow cell (FLO-MIN114), and sequencing was carried out for 48 h.

Basecalling was performed using Dorado basecall server Version 7.3.9 in super-accuracy mode. The expected genome size was set to 5.1 Mb for coverage estimation and downstream assembly. Basecalled ONT reads were quality filtered using Filtlong v0.2.1 [22]; reads shorter than 1 kb and the lowest-scoring 5% of read bases were discarded. Only samples retaining ≥100× filtered-read coverage, calculated as total filtered bases divided by the expected genome size of 5.1 Mb, were included for assembly. Filtered reads were de novo assembled using Flye (v2.9.6) [23]. Draft assemblies were polished with one round of Racon v1.5.0 [24] after mapping filtered reads back to the assembly, followed by one round of Medaka v1.5.0 [25]. Contigs shorter than 500bp were discarded. Assembly completeness and contamination were evaluated using CheckM [26].

Following assembly and quality control, Sequence types were determined by MLST v.2.33.0 [27, 28]. Antimicrobial resistance genes (ARGs) were identified using AMRFinderPlus v3.10.30 [29]. Plasmid typing was performed using MOB-suite v3.1.9 [30] and PubMLST to characterize the plasmid content of the CR-*E. coli* isolates [27]. For plasmid typing via PubMLST, plasmids with less than 100% locus matches known replicon types were categorized as “No possible id”. Finally, a Sankey diagram was constructed to visualize the interrelationships from clinical specimen type to sequence type, carbapenemase gene, and plasmid type.

### 2.5. Core-genome SNP phylogenetic analysis

Allele profiles for core-genome multilocus sequence typing (cgMLST) were determined using the EnteroBase *Escherichia coli* cgMLST v1 scheme (comprising 2,513 core loci) [31]. A reference pseudogenome was composed using all the cgMLST genes, and sequencing reads from each isolate were aligned against this pseudogenome using Snippy v4.6.0 [32]. Core-genome single-nucleotide polymorphisms (SNPs) were extracted from the resulting alignments using snippy-core. Only single-copy core alleles shared between the reference and study isolates were retained for SNP profiling.

Pairwise SNP distance matrices were generated using snp-dists v0.8.2 to assess the genetic relatedness among isolates [33]. Maximum-likelihood phylogenetic trees were inferred from the core SNP alignment using IQ-TREE v2.4.0 with 1,000 bootstrap replicates, using the GTR+F+I+G substitution model as the best-fit model determined by ModelFinder [34, 35].

Given that cross-sectoral dissemination along the human–food interface involves variable temporal windows, intermediate vectors, and diverse selection pressures, pairwise SNP distances were evaluated using an operational framework informed by established One Health genomic surveillance parameters for *E. coli* [36, 37]. To maintain high phylogenetic stringency while capturing cross-sectoral relatedness otherwise obscured by acute outbreak metrics, a threshold of ≤50 SNPs was established as the primary cutoff for defining closely related cross-sectoral genomic clusters. Pairwise distances of 51–100 SNPs were further evaluated to identify broader cross-sectoral linkages indicative of shared upstream reservoirs [36, 37]. Phylogenetic trees were midpoint-rooted and annotated with isolate metadata using the Interactive Tree Of Life (iTOL) [38].

### 2.6. Comparative genomic and plasmid analysis

Comparative genomic analysis was conducted on a major phylogenetic clade of interest; that clade was selected due to its inclusion of highly related isolates from diverse sources, highlighting potential cross-sectoral genomic overlap. To evaluate whole-genome relatedness within this phylogenetic clade, average nucleotide identity (ANI) analysis was conducted using ANIclustermap [39].

To investigate the genetic environment and structural conservation of *bla*_NDM-5_-carrying plasmids, the complete plasmid sequence pEC-16-10-NDM-5 from *Escherichia coli* strain EC-16-10 (GenBank accession no. MZ836801.1) was selected as a reference [40]. This plasmid was selected for accurate structural mapping and comparison with the plasmids identified in our study isolates. The reference plasmid was annotated using the Proksee server to identify key plasmid-associated features, including antimicrobial resistance genes, transposase-related elements, and conjugative transfer machinery [41].

For detailed structural comparison, eight representative ST69 isolates from this phylogenetic clade were selected, comprising two isolates each from raw beef, raw chicken, raw pork, and clinical sources. The plasmid contigs of these representative isolates were aligned and visualized against the reference plasmid using Easyfig (v2.2.5) to assess local rearrangements, gene relocations, and synteny [42].

## 3. Results

### 3.1. Characterization of clinical CR-*Escherichia coli*

Among the 271 clinical CR-*E. coli* isolates, rectal screening swabs represented the predominant specimen source, accounting for 78.2% (212/271) of isolates, followed by urine (10.7%, 29/271), stool (6.3%, 17/271), blood (1.5%, 4/271), wound swabs (1.5%, 4/271), and other clinical specimen types (1.8%, 5/271) (Figure 2A). MLST revealed that ST69 was the most prevalent sequence type (ST), comprising 13.3% (36/271) of the isolates. Other recurrent sequence types included ST10 (5.2%, 14/271), ST101 (4.4%, 12/271), ST155 (3.3%, 9/271), ST131 (3.0%, 8/271), ST48 (3.0%, 8/271), ST744 (3.0%, 8/271), ST167 (2.6%, 7/271), ST3580 (2.6%, 7/271), and ST93 (2.6%, 7/271). Of the remaining isolates, 8.5% (23/271) corresponded to unassigned sequence types, while 48.7% (132/271) were distributed across a highly diverse population of low-frequency sequence types containing 1 to 5 isolates each (<2.0% of the clinical collection per ST).

**Figure 2:**
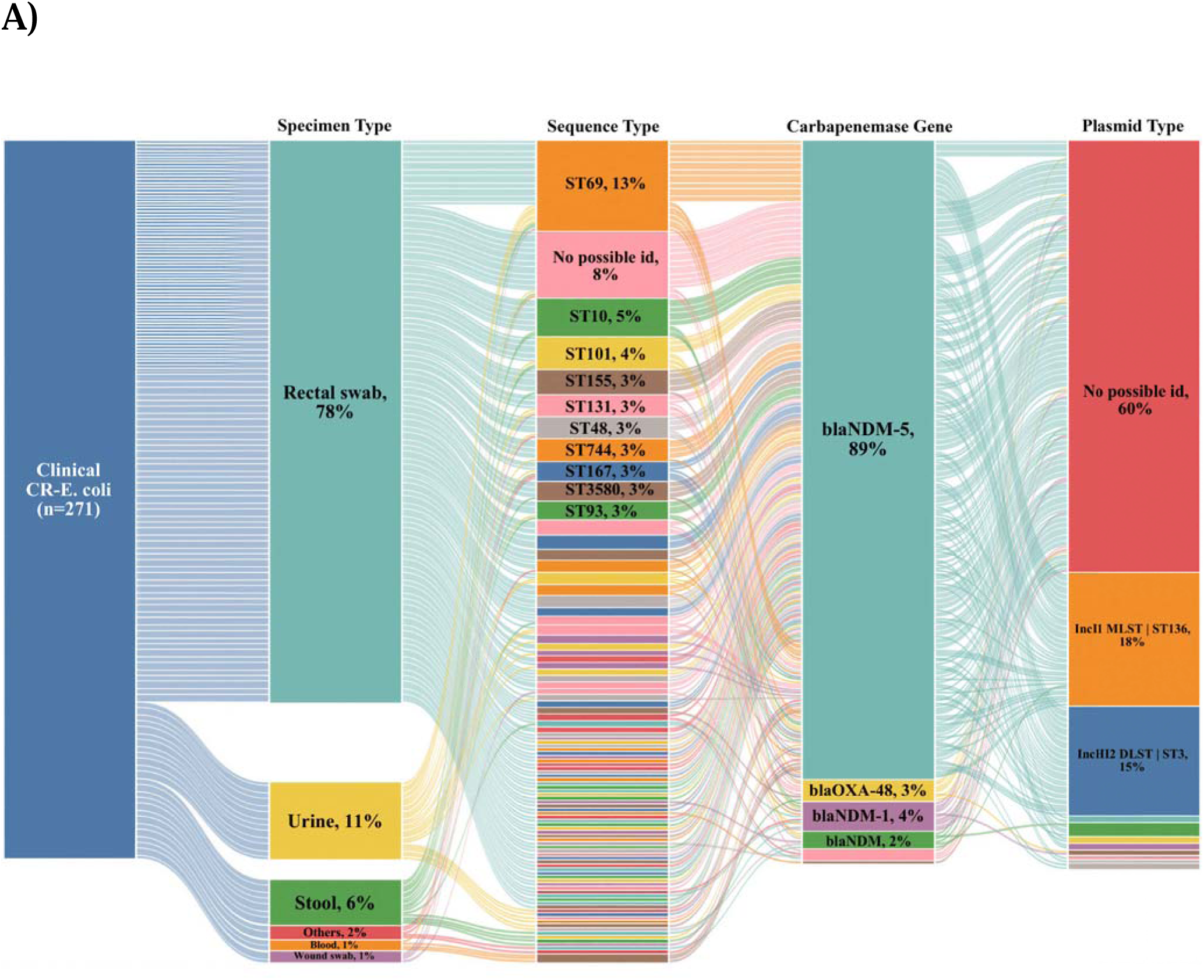
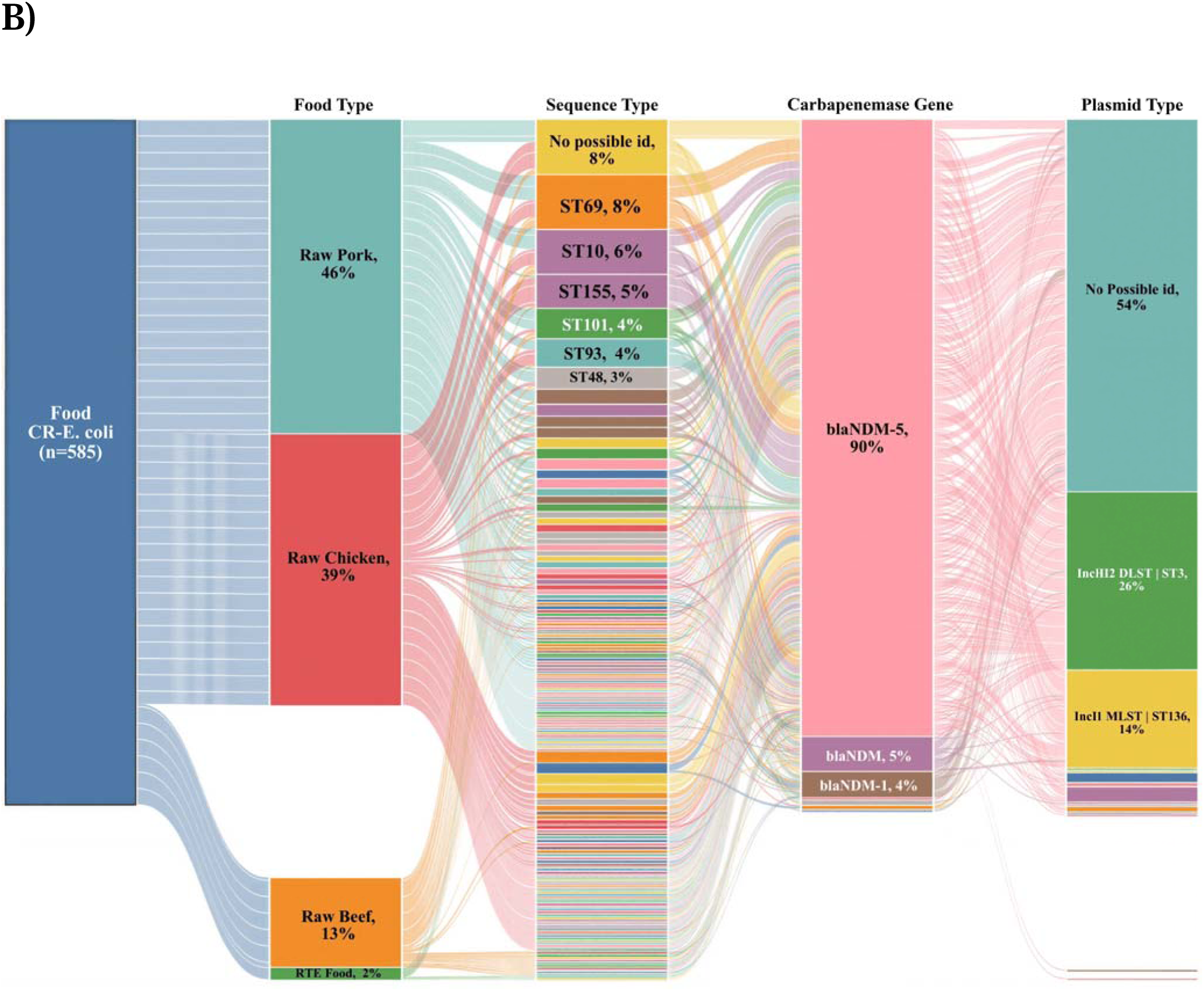
Sankey diagrams showing the association among sample types, sequence types (STs), carbapenemase genes, and plasmid types for carbapenem-resistant *Escherichia coli* (CR-*E. coli*) isolates from (A) clinical specimens and (B) food samples. The width of each band is proportional to the number of isolates represented in each category.

Carbapenemase gene profiling demonstrated that *bla*_NDM-5_ was the primary driver of carbapenem resistance, detected in 88.9% (241/271) of clinical isolates. This resistance gene was broadly disseminated across diverse sequence-type backgrounds, including the predominant ST69 lineage. Other carbapenemase determinants were detected sporadically, including *bla*_NDM-1_ (4.1%, 11/271), *bla*_OXA-48_ (3.0%, 8/271), *bla*_OXA-181_ (1.5%, 4/271), and unsubtyped *bla*_NDM_ (2.2%, 6/271), with one isolate (0.4%) co-harboring *bla*_NDM-5_ and *bla*_OXA-181_.

Plasmid replicon typing revealed that IncI1 MLST | ST136 (18.5%, 50/271) and IncHI2 DLST | ST3 (15.1%, 41/271) represented the most frequent typable plasmid profiles associated with this clinical collection. Less common typable replicon backbones included IncI1 MLST | ST80 (1.8%, 5/271), IncHI2 DLST | ST2 (0.7%, 2/271), IncI1 MLST | ST16 (0.7%, 2/271), IncHI1 MLST | ST17 (0.7%, 2/271), IncI1 MLST | ST162 (0.7%, 2/271), IncI1 MLST | ST20 (0.7%, 2/271), IncI1 MLST | ST167 (0.4%, 1/271), and IncI1 MLST | ST223 (0.4%, 1/271). A total of 163 isolates (60.1%) harbored untypable plasmid replicons or lacked defined replicons present in the reference database.

### 3.2. Characterization of food-derived CR-*Escherichia coli*

A total of 585 food-derived CR-*E. coli* isolates were recovered from 4,917 retail food samples, representing an overall isolate recovery rate of 11.9%. Raw pork constituted the primary food reservoir, accounting for 45.8% (268/585) of isolates, followed by raw chicken (39.5%, 231/585) and raw beef (13.0%, 76/585). Ready-to-eat (RTE) foods contributed a minority of isolates (1.7%, 10/585) (Figure 2B).

MLST analysis demonstrated substantial clonal diversity among the foodborne CR-*E. coli* isolates. Mirroring the clinical collection, ST69 emerged as the single most common sequence type, comprising 7.5% (44/585) of the food-derived isolates. Other prominent sequence types included ST10 (6.5%, 38/585), ST155 (4.8%, 28/585), ST101 (4.3%, 25/585), ST93 (3.9%, 23/585), and ST48 (3.1%, 18/585). Unassigned sequence types accounted for 8.2% (48/585) of isolates, while the remaining 361 isolates (61.7%) were distributed across a broad spectrum of low-frequency sequence types containing 1 to 12 isolates each (≤2.0% per ST).

Carbapenemase gene profiling established that *bla*_NDM-5_ was the overwhelming carbapenemase determinant in the food sector, identified in 90.1% (527/585) of isolates. Similar to clinical observations, *bla*_NDM-5_ was distributed across multiple clonal backgrounds, led by ST69. Additional resistance gene profiles were observed at lower frequencies, including un-subtyped *bla*_NDM_ (5.0%, 29/585), *bla*_NDM-1_ (3.8%, 22/585), co-carriage of *bla*_NDM-5_ and *bla*_OXA-181_ (0.5%, 3/585), co-carriage of *bla*_NDM-1_ and *bla*_NDM-5_ (0.5%, 3/585), and *bla*_OXA-181_ alone (0.2%, 1/585).

Plasmid replicon typing identified IncHI2 | DLST ST3 as the most prevalent typable plasmid profile among foodborne isolates, present in 25.8% (151/585) of the collection, followed closely by IncI1 MLST | ST136 in 14.2% (83/585) of isolates. Minor typable plasmid profiles included IncI1 MLST | ST80 (2.2%, 13/585), IncHI2 DLST | ST2 (1.4%, 8/585), IncHI1 MLST | ST17 (0.5%, 3/585), IncI1 MLST | ST162 (0.5%, 3/585), IncI1 MLST | ST271 (0.2%, 1/585%), IncI1 MLST | ST113 (0.2%, 1/585%), IncI1 MLST | ST13 (0.2%, 1/585%), IncI1 MLST | ST167 (0.2%, 1/585%), IncI1 MLST | ST181 (0.2%, 1/585%), IncI1 MLST | ST183 (0.2%, 1/585%), and IncI1 MLST | ST21 (0.2%, 1/585%). Over half of the foodborne isolates (54.2%, 317/585) harbored untypable plasmid replicons or lacked defined reference replicon matches.

### 3.3. Comparison of sequence type distributions between clinical and food-derived CR-*Escherichia coli*

By performing a merged MLST distribution across both clinical and food sources, the five most prevalent sequence types in the combined dataset were ST69 (n=80; 36 clinical, 44 food-derived), ST10 (n=52; 14 clinical, 38 food-derived), ST155 (n=37; 9 clinical, 28 food-derived), ST101 (n=37; 12 clinical, 25 food-derived), and ST93 (n=30; 7 clinical, 23 food-derived) (Figure 3). Because ST69 was the most predominant defined sequence type shared across both clinical and food sources, it was selected for subsequent phylogenetic, comparative genomic and plasmid analyses.

**Figure 3:**
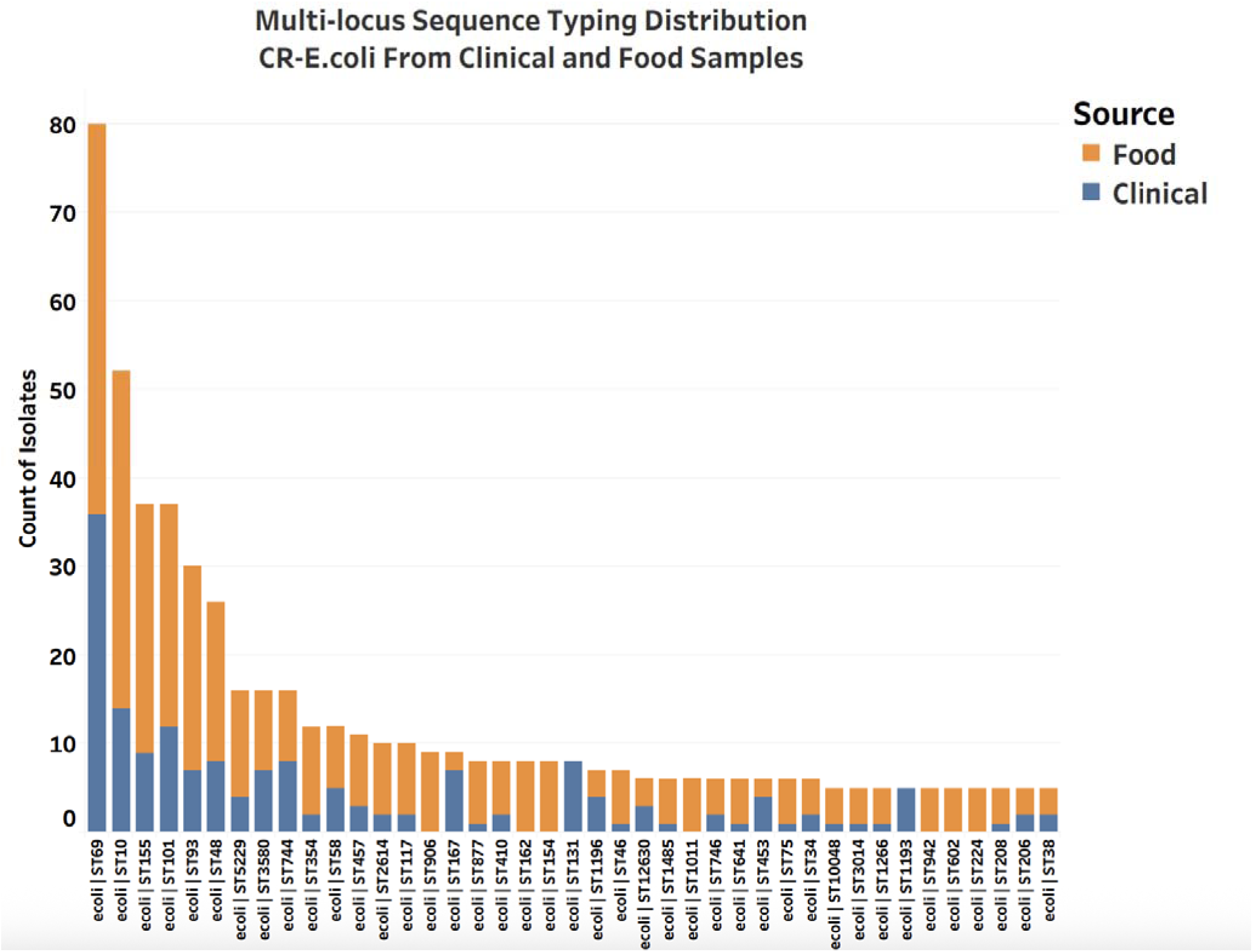
Multilocus sequence typing (MLST) distribution of carbapenem-resistant *Escherichia coli* (CR-*E. coli*) isolates from clinical and food sources. For visual clarity, low-frequency sequence types (STs) represented by four or fewer isolates (≤4) were excluded.

### 3.4. Core-genome SNP phylogenomics of ST69 isolates across clinical and food sectors

To evaluate the genetic relatedness between clinical and food-derived ST69 CR-*E. coli* isolates, a maximum-likelihood phylogenetic tree based on core-genome single-nucleotide polymorphisms (SNPs) was constructed. Applying the primary operational threshold of ≤ 50 core SNPs, ST69 isolates were partitioned into distinct phylogenetic groupings, centered on a major mixed-source lineage designated Clade 1, alongside smaller mono-source clusters (Figure 4).

**Figure 4.**
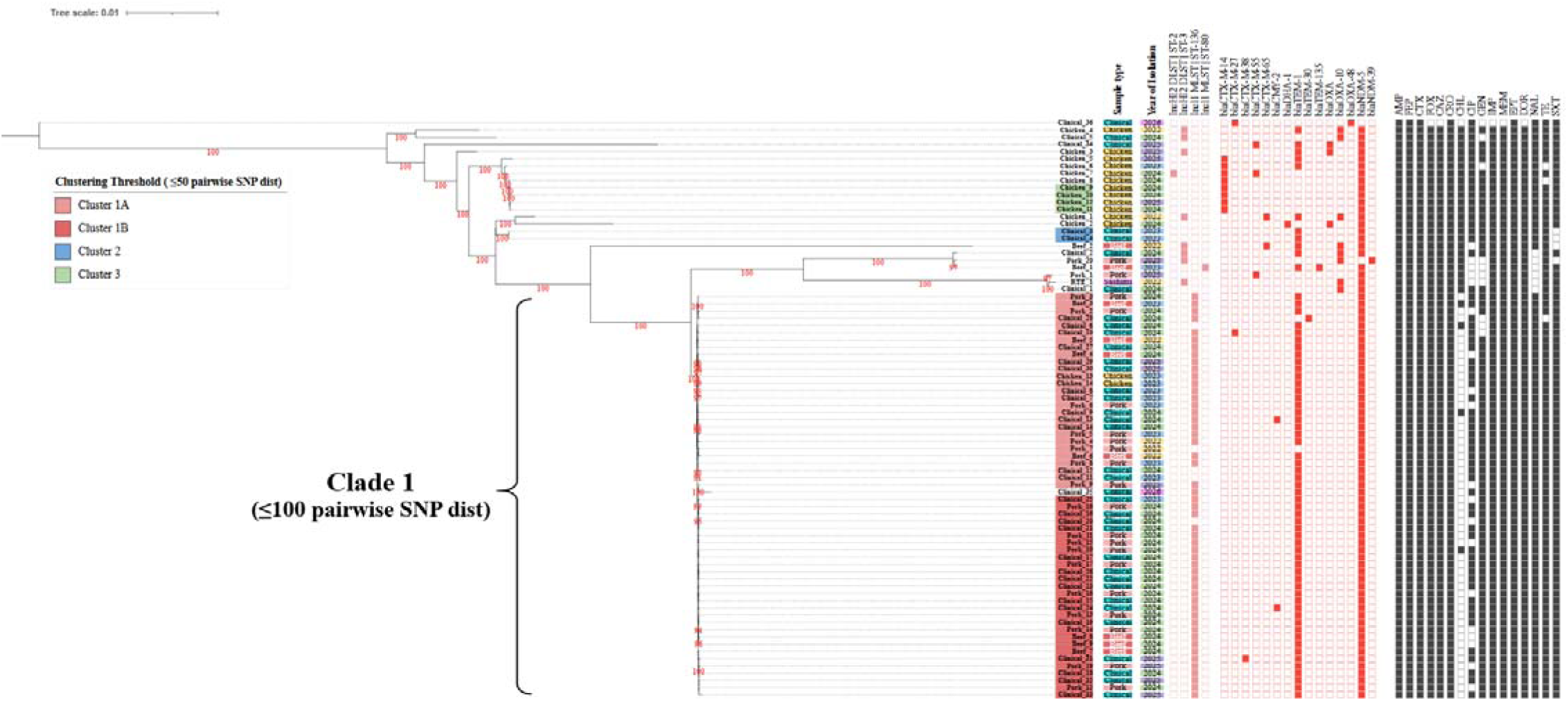
Phylogenetic analysis of ST69 CR-*E. coli* isolates from clinical and food sources based on core-genome single-nucleotide polymorphisms. Closely related isolates were defined using a predefined ≤50-SNP threshold. Clusters 1A and 1B represent major mixed-source clusters comprising clinical and food-derived isolates (Tree nodes highlighted in red). Cluster 2 represents a clinical-specific lineage (Tree nodes highlighted in blue), and Cluster 3 represents a food-specific lineage recovered from retail chicken samples (Tree nodes highlighted in green). Clinical_35 was positioned within the broader exploratory Clade 1 under the 51–100-SNP window but was not included in the ≤50-SNP clusters.

At the predefined ≤50-SNP threshold, 55 ST69 isolates formed two major mixed-source clusters, Clusters 1A and 1B, comprising 28 clinical and 27 food-derived isolates. Within these two clusters, internal pairwise SNP distances ranged from 0 to 31 SNPs, indicating close genomic relatedness among isolates recovered from both sectors. One additional clinical isolate, Clinical_35, was separated from these clusters by 51–100 SNPs and was therefore included only in the broader exploratory Clade 1, which comprised 56 isolates in total, including 29 clinical and 27 food-derived isolates (Figure 4).

Isolates within Clusters 1A and 1B exhibited highly concordant multidrug-resistant phenotypic profiles across the antimicrobials tested. All isolates displayed uniform non-susceptibility to penicillins (ampicillin), broad-spectrum cephalosporins/cephamycins (cefepime, cefotaxime, cefoxitin, ceftazidime, and ceftriaxone), carbapenems (imipenem, meropenem, ertapenem, and doripenem), nalidixic acid, tetracyclines (tetracycline), and folate pathway antagonists (trimethoprim-sulfamethoxazole). Phenotypic variation was restricted to chloramphenicol, ciprofloxacin, and gentamicin. Genotypically, isolates in Clusters 1A and 1B shared a conserved resistome and plasmid architecture, characterized by the co-carriage of *bla*_NDM-5_ and *bla*_TEM-1_ on IncI1 MLST | ST136 plasmids.

Cluster 2 represented a clinical-specific lineage comprising two ST69 CR-*E. coli* isolates recovered from two patients in 2023 (blue tree nodes, Figure 4). These isolates exhibited zero SNP differences (0 SNPs) and displayed identical antimicrobial susceptibility profiles, demonstrating non-susceptibility to all tested agents except trimethoprim-sulfamethoxazole. Furthermore, both Cluster 2 isolates co-harbored *bla*_NDM-5_ and *bla*_TEM-1_.

Cluster 3 represented a food-specific lineage comprising four ST69 CR-*E. coli* isolates recovered exclusively from retail chicken samples across 2024 and 2025 (green tree nodes, Figure 4). Pairwise distances within Cluster 3 ranged from 10 to 12 SNPs. All four isolates exhibited complete non-susceptibility to all tested antimicrobial agents. Notably, in contrast to Clusters 1A, 1B and 2, all isolates in Cluster 3 co-carried *bla*_CTX-M-14_ and *bla*_NDM-5_.

### 3.5. Whole-genome identity and comparative plasmid synteny of Clade 1 isolates

Comparative genomic analysis of the 56 ST69 CR-*E. coli* isolates in Clade 1 showed pairwise whole-genome average nucleotide identity (ANI) values ranging between 99.85% and 100.00% (Figure 5). Hierarchical clustering based on ANI values showed that clinical isolates and food-derived isolates (raw beef, raw pork, and raw chicken) were distributed across shared branches of the ANI heatmap and dendrogram (Figure 5).

**Figure 5:**
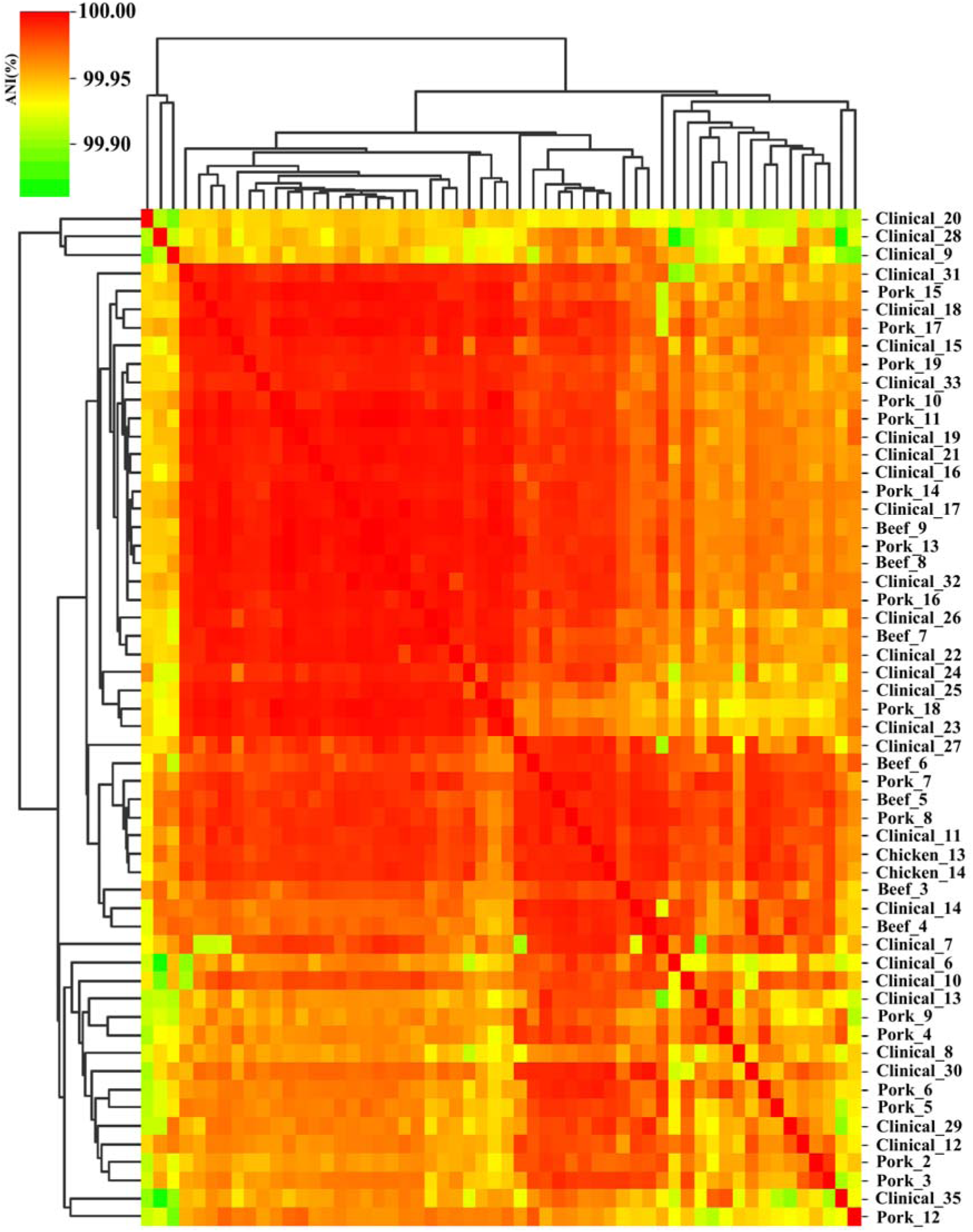
Whole-genome comparative analysis of clustered ST69 CR-*Escherichia coli* isolates. Heatmap depicting Average Nucleotide Identity (ANI)-based hierarchical clustering of 56 ST69 carbapenem-resistant *Escherichia coli* (CR-*E. coli*) isolates belonging to phylogenetic Clade 1 (≤100 core-genome SNP differences). The matrix illustrates a high degree of whole-genome similarity (>99.85%) among isolates recovered across clinical and food sources.

The genetic environment of *bla*_NDM-5_ and the plasmid backbone structure were evaluated relative to the reference IncI1 plasmid pEC-16-10-NDM-5 (GenBank accession: MZ836801.1). Circular genetic mapping (Proksee) identified the *bla*_NDM-5_ gene adjacent to dsbC, umuD, umuC, parB, insertion element insK, relaxase gene, and conjugative transfer/pilus assembly operons (tra and pil) (Figure 6A).

**Figure 6.**
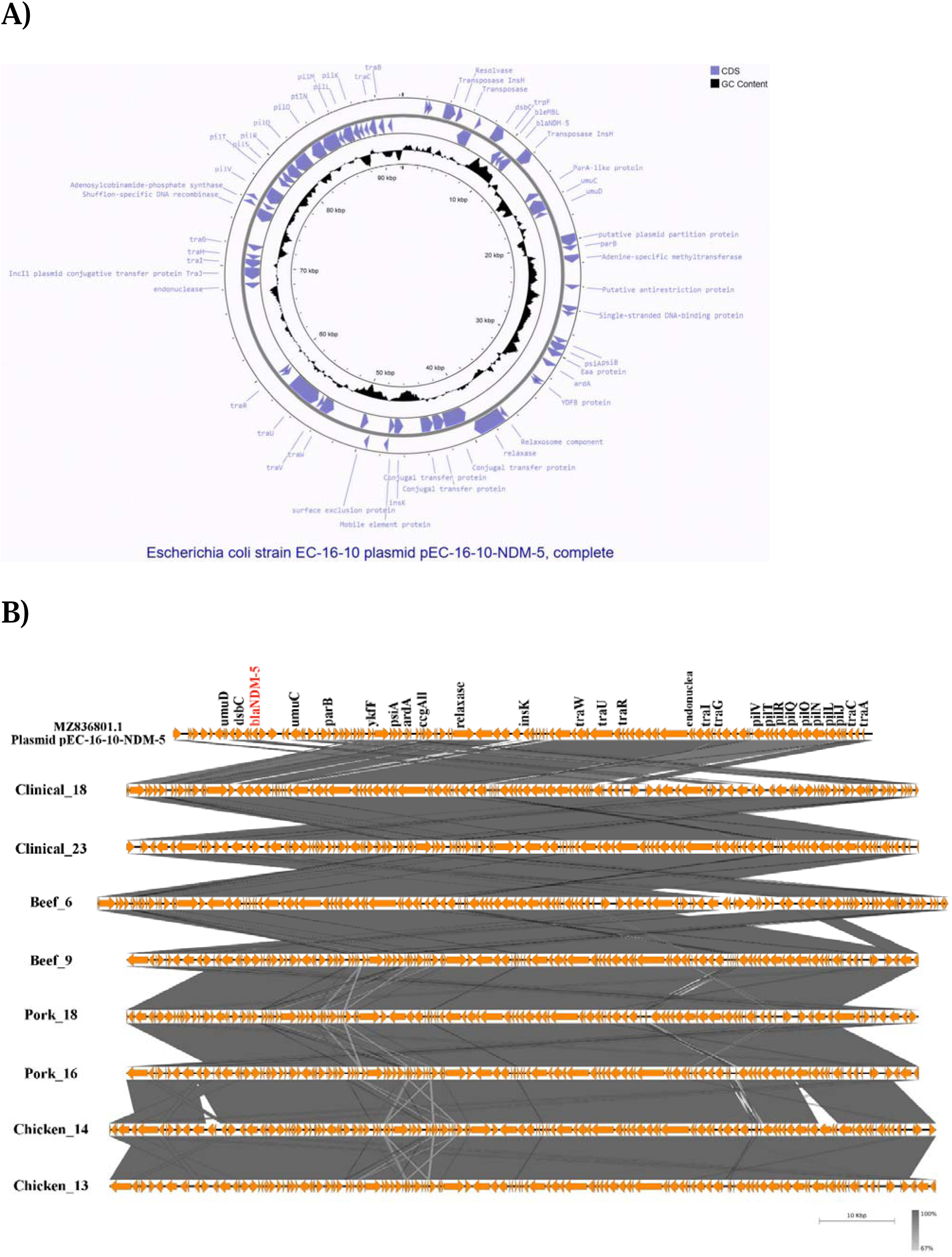
Comparative analysis of the *bla*_NDM−5_ carrying plasmid associated with clustered ST69 CR-*Escherichia coli* isolates. **(A) Proksee map of the reference plasmid pEC-16-10-NDM-5 from *Escherichia coli* strain EC-16-10 (GenBank accession no. MZ836801.1),** showing the annotated plasmid structure and key genetic features. **(B) Easyfig alignment of pEC-16-10-NDM-5 (MZ836801.1) with plasmid contigs from eight representative ST69 CR-*E. coli* isolates, comprising two isolates each from raw beef, raw chicken, raw pork, and clinical sources.** Shaded regions indicate homologous sequences between plasmids.

Linear plasmid alignment (Easyfig) was performed across eight representative ST69 isolates, comprising two isolates each from raw beef, raw chicken, raw pork, and clinical specimens (Figure 6B). The aligned plasmid contigs displayed sequence synteny across the *bla*_NDM-5_-carrying IncI1 backbone among all four source types, with pairwise BLAST nucleotide identity ranging from 67% to 100% (dark grey shading indicating >90% identity across core regions). Sequence variation and local structural rearrangements were observed primarily in regions associated with insertion sequence insK and within conjugative transfer regions containing tra (traI, traG, traC, traW, traU, traR) and pil (pilV, pilT, pilR, pilQ, pilO, pilN, pilL, pilJ) genes.

## 4. Discussion

The present study identified a marked shift in the molecular epidemiology of CR-*E. coli* in Hong Kong. ST69 was the most frequent defined sequence type among both clinical isolates (36/271, 13.3%) and food-derived isolates (44/585, 7.5%). Its prominence is notable relative to a previous local study in which ST69 was not detected among clinical CPE isolates [19]. This apparent emergence occurred against a background of widespread *bla*_NDM-5_ dissemination across diverse sequence types. Moreover, a subset of clinical and food-derived ST69 isolates formed a closely related mixed-source phylogenetic cluster, demonstrated >99.85% average nucleotide identity, and was associated with similar *bla*_NDM-5_-bearing IncI1 plasmid profiles. To our knowledge, this study provides initial genomic characterization of cross-sectoral overlap between clinical and food-derived CR-*E. coli* isolates in Hong Kong.

The predominance of *bla*_NDM-5_ in the clinical collection is clinically relevant because CPE are frequently multidrug resistant and associated with limited treatment options [1–6,9]. However, most clinical isolates in this study were recovered from rectal-screening specimens. The findings should therefore be interpreted primarily as evidence of gastrointestinal carriage rather than invasive infection. This distinction is important, but carriage remains epidemiologically meaningful, as the intestinal tract can serve as a reservoir for subsequent infection, persistence within healthcare settings, and onward dissemination.

Compared with a previous genomic study of clinical CPE isolates in Hong Kong hospitals collected between 2011 and 2017 [19], the present collection exhibited a higher representation of ST69 and a pronounced dominance of *bla*_NDM-5_. Although the studies were not designed as a formal longitudinal surveillance series, the contrast is informative. ST69 was not detected in that earlier clinical dataset [19], whereas it represented the most frequent defined ST in both the current clinical and food-derived collections. This supports an important contemporary shift in the sampled CR-*E. coli* population, while formal confirmation of lineage replacement would require harmonized longitudinal surveillance across comparable sources and time periods.

The recovery of closely related ST69 isolates from both clinical carriage and retail meat places these findings in a One Health context. Official raw-meat AMR bacteria surveillance in Hong Kong has repeatedly detected CPE in retail food samples, indicating that raw meat is a recurring reservoir of resistant organisms [16]. The present study extends this observation by showing that a clinically relevant CR-*E. coli* lineage was present in both clinical and food-derived collections. These findings do not demonstrate direct food-to-human transmission, but they indicate that raw retail meat is an important interface for monitoring the circulation of carbapenemase-producing *E. coli*.

The mixed-source ST69 cluster provides the strongest evidence for cross-sector genomic overlap. Clinical and food-derived isolates clustered closely by core-genome SNP analysis, and ANI analysis demonstrated >99.85% nucleotide identity, supporting their assignment to a highly similar genomic population. However, there is currently no universal SNP threshold for defining clonality or recent transmission in CR-*E. coli* across different hosts, sources, and sampling intervals. In this study, isolates within ≤50 SNPs were considered a stringent signal of close relatedness based on previous literature, whereas the 51–100 SNP range was used as a broader discovery window for One Health cross-sector linkage [36, 37]. This wider range is useful in food-chain investigations, where related isolates may be separated by time, unsampled intermediates, supply-chain movement, or diversification during colonization and environmental persistence. Therefore, the ≤50-SNP cluster supports close genetic relatedness, while the 51–100 SNP range helps identify a wider related population that may otherwise be missed by overly restrictive thresholds.

Plasmid comparisons provide additional context for this cross-sectoral signal. Representative clinical and food-derived ST69 isolates carried similar IncI1 plasmid profiles associated with *bla*_NDM-5_. Conservation of the *bla*_NDM-5_ region and plasmid backbone suggests that related mobile elements contributed to the observed cross-sector signal. At the same time, structural variation around mobile genetic elements and transfer-associated regions suggests structural divergence in local plasmid backbones. These data are consistent with the presence of related plasmid backbones across bacterial populations, although functional conjugation assays and complete long-read plasmid sequencing would be required to evaluate horizontal plasmid transfer dynamics.

This IncI1-associated signal differs from those reported in mainland China and other settings, where *bla*_NDM-5_ dissemination in food animals and retail meat has frequently been linked to IncX3 plasmids [17,18,38,43]. The present findings therefore suggest that the Hong Kong ST69 cluster may not be explained solely by the IncX3-driven dissemination pattern described elsewhere. Instead, expansion of a successful ST69 background, maintenance of a related IncI1 *bla*_NDM-5_ plasmid, or both may have contributed. Previous reports linking ST69 to human infection and retail meat, together with reports of IncI1-associated carbapenemase carriage in avian *E. coli*, support the biological plausibility of this lineage–plasmid combination [44–46].

Whether the observed genomic overlap originated through upstream importation, local amplification, or a combination of both remains uncertain. Hong Kong relies heavily on imported food, including fresh meat, and mainland China remains an important supplier [47–49], suggesting that contamination may arise upstream before entering the local market. However, studies from mainland China have more frequently highlighted ST48, ST101, ST156, ST10, and ST155 in food-associated multidrug-resistant *E. coli*, rather than ST69 [50–53]. This discrepancy raises the possibility that local processing in Hong Kong, including slaughtering, distribution, and retail handling, provides critical opportunities for local amplification and cross-contamination of ST69. The persistence of related ST69 isolates across different sources and years is compatible with ongoing circulation or repeated introduction somewhere along the supply chain, but more detailed source-attribution data are needed to distinguish between these scenarios.

Several limitations of the study should be acknowledged. First, genomic relatedness demonstrates co-circulation but cannot establish the exact directionality of transmission between food and human sources. Second, the clinical collection was dominated by rectal-screening isolates from four hospitals and should not be interpreted as demonstrating a corresponding increase in the burden of CR-*E. coli* disease. Third, detailed phylogenetic and plasmid analyses focused on ST69. Genomic relationships involving other shared lineages may therefore have been overlooked. Fourth, there is currently no universal consensus on the exact SNP threshold for defining clonal relatedness of CR-*E. coli*, although this study utilized a ≤50 SNP threshold based on previous literature [36, 37]. Finally, the absence of isolates from food-producing animals, slaughter and processing environments, community populations, and wastewater limited source attribution and the identification of intermediate reservoirs. Despite these limitations, the convergence of phenotypic, phylogenetic, and plasmid-associated findings identifies *bla*_NDM-5_-carrying ST69 as a newly prominent and epidemiologically important CR-*E. coli* lineage in Hong Kong and supports continued integrated One Health surveillance.

## 5. Conclusion

This study provides updated molecular epidemiological evidence of a shift in CR-*E. coli* populations in Hong Kong, with *bla*_NDM-5_-bearing ST69 emerging as the most frequent defined sequence type in both clinical and food-derived collections. Genomic analyses showed that isolates from the two sectors were closely related, exhibited exceptional whole-genome similarity within the mixed-source ST69 cluster, and carried highly similar *bla*_NDM-5_-associated IncI1 plasmid profiles. These findings support the possibility of co-circulation between human and food-associated reservoirs, although direct transmission and its directionality cannot be inferred from the available data.

The predominance of related isolates from raw pork and chicken highlights the food chain as a potential exposure interface for CR-*E. coli* in Hong Kong. Together, these data support a One Health perspective on the emergence and spread of CR-*E. coli* locally. Continued integrated surveillance across clinical, food, animal, and environmental sectors, together with complete plasmid characterization and source-tracing studies, will be important to clarify transmission pathways and inform targeted containment strategies.

## Ethics Statement

This study was approved by the Institutional Review Boards (IRB) of The Hong Kong Polytechnic University (ARSA-23248-HMRF-HTI). The current study collected clinical isolates of Carbapenem-Resistant *Escherichia coli* from public hospitals under the approval of the Hospital Authority Central IRB (ref. no. CIRB-2023-222-3).

## Data availability

Raw sequencing data for all ST69 CR-*E. coli* isolates are available in the NCBI Sequence Read Archive (SRA) under BioProject PRJNA1282312.

